# A confounder debiasing method for RCT-like comparability enables Machine Learning-based personalization of survival benefit in living donor liver transplantation

**DOI:** 10.1101/2024.11.01.24316601

**Authors:** Anirudh Gangadhar, Bima J. Hasjim, Xun Zhao, Yingji Sun, Joseph Chon, Aman Sidhu, Elmar Jaeckel, Nazia Selzner, Mark S. Cattral, Blayne A. Sayed, Michael Brudno, Chris McIntosh, Mamatha Bhat

## Abstract

Many clinical questions in medicine cannot be answered through randomized controlled trials (RCTs) due to ethical or feasibility constraints. In such cases, observational data is often the only available resource for evaluating treatment effects. To address this challenge, we have developed Decision Path Similarity Matching (DPSM), a novel machine learning (ML)-based algorithm that simulates RCT-like conditions to debias observational data. In this study, we apply DPSM to the clinical question of living donor liver transplantation (LDLT) versus deceased donor liver transplantation (DDLT), helping to identify which patients benefit most from LDLT. DPSM leverages decision paths from a Random Forest classifier to perform accurate, one-to-one matching between LDLT and DDLT recipients, minimizing confounding while retaining interpretability. Using data from the Scientific Registry of Transplant Recipients (SRTR), including 4,473 LDLT and 68,108 DDLT patients transplanted between 2002 and 2023, we trained independent Random Survival Forest (RSF) models on the matched cohorts to predict post-transplant survival. DPSM successfully reduced confounding associations between the two groups as shown by a decrease in area under the receiver operating characteristic (AUROC) from 0.82 to 0.51. Subsequently, RSF (C-index_ldlt_=0.67, C-index_ddlt_=0.74) outperformed the traditional Cox model (C-index_ldlt_=0.57, C-index_ddlt_=0.65). The predicted 10-year mean survival gain was 10.3% (SD = 5.7%). In conclusion, DPSM provides an effective approach for creating RCT-like comparability from observational data, enabling personalized survival predictions. By leveraging real-world data where RCTs are impractical, this method offers clinicians a tool for transitioning from population-level evidence to more nuanced, personalization.

## 1. Main

Many clinical questions in medicine cannot be addressed through randomized controlled trials^1^ (RCTs) due to ethical, logistical, or practical challenges. In liver transplantation (LT), for example, it is not feasible to randomize patients between living donor liver transplantation (LDLT) and deceased donor liver transplantation (DDLT) because of the ethical implications of assigning healthy donors and the urgency for life-saving transplants^2–4^. As a result, we often rely on observational data to assess the relative benefits of LDLT and DDLT.

Clinically, LDLT offers significant advantages over DDLT, such as reduced waitlist times^5–8^, improved graft quality^5,9^, and lower rejection rates^10^. Despite these benefits, LDLT remains underutilized, representing only 5% of all LT cases in the United States^6^. Previous studies^5,10,11^ have shown general survival benefits of LDLT compared to DDLT, but they lack sufficient adjustment for confounding factors, making it difficult for clinicians to determine which individual patients would benefit most from LDLT.

RCTs are considered the gold standard for assessing intervention effects, but as mentioned, they are not feasible in the LDLT versus DDLT context. This has led to the development of advanced statistical and machine learning methods that can simulate RCT-like conditions using observational data. Propensity Score Matching (PSM)^12,13^ is one such method, but it has limitations. By reducing complex, multi-dimensional covariate space into a single probability score, PSM can fail to balance key variables and interactions, leading to residual confounding and imprecise graft-type effect estimates^14–16^.

In response to these limitations, we introduce Decision Path Similarity Matching (DPSM), a novel machine learning-based algorithm designed to improve matching by leveraging the decision paths from Random Forest models. Unlike PSM, our method matches patients based on entire decision paths rather than a single probability score. This richer representation captures complex, non-linear relationships between covariates, enabling more precise matching and minimizing confounding. DPSM also allows for explainability by providing per-matched-pair visualization of the key variables driving the decision-making process.

After applying DPSM to match LDLT and DDLT patients, we utilize a time-to-event machine learning framework, specifically Random Survival Forest (RSF) models, to predict long-term survival outcomes. To the best of our knowledge, no existing method offers individualized survival benefit predictions for LDLT versus DDLT based on patient-specific variables, making this work a significant advancement in the field.

## 2. Results

### 2.1. Patient characteristics

A total of 72,581 LT recipients were included in the study. DDLTs constituted 93.8% (*n*_*ddlt*_ = 68,108), while LDLTs comprised a much lesser percentage at 6.2%, (*n*_*ldlt*_ = 4,473). Demographic and clinical study variables for both groups are reported in Table 1. DDLT patients had higher rates of post-transplant mortality (29.7%) as compared to LDLT (22.3%).

**Table 1.** Clinicodemographic characteristics of LDLT, DDLT recipients.

| Characteristics | LDLT population<br>(n = 4,473) | DDLT population<br>(n = 68,108) | p-value |
| --- | --- | --- | --- |
| Age (years) | 51.5 (12.8) | 52.9 (10.4) | <0.001 |
| Sex |  |  |  |
| Female (%) | 48.3 | 35.6 |  |
| Male (%) | 51.7 | 64.4 | <0.001 |
| Height (cm) | 169.9 (10.3) | 172.3 (10.3) | <0.001 |
| Weight (kg) | 78.8 (17.7) | 87.2 (20.3) | <0.001 |
| BMI | 27.2 (5.2) | 29.3 (5.9) | <0.001 |
| Blood type (%) |  |  |  |
| A | 43.3 | 36.7 | <0.001 |
| AB | 1.9 | 5.4 | <0.001 |
| B | 9.8 | 13.5 | <0.001 |
| O | 45.0 | 44.4 | 0.375 |
| MELD | 14.0 (4.5) | 22.4 (8.7) | 0.000 |
| Primary etiology (%) |  |  |  |
| Alcoholic Cirrhosis | 15.1 | 29.4 | <0.001 |
| Autoimmune Hepatitis | 4.6 | 3.5 | <0.001 |
| Cryptogenic Cirrhosis | 6.9 | 6.6 | 0.378 |
| HBV Cirrhosis | 1.0 | 1.7 | <0.001 |
| HCV Cirrhosis | 15.8 | 22.1 | <0.001 |
| Metabolic Liver Disease | 2.0 | 2.2 | 0.374 |
| MASH | 16.9 | 15.3 | 0.007 |
| PSC | 19.0 | 5.2 | 0.956 |
| PBC | 8.9 | 3.7 | <0.001 |
| Other | 8.0 | 7.9 | <0.001 |

### 2.1. LDLT-DDLT matching

In our study, where the goal is to estimate the survival benefit associated with receiving one type of intervention (LDLT) over another (DDLT), it becomes necessary to minimize confounding associations to ensure that our findings are not subject to bias. To this end, we developed the DPSM matching algorithm (details in Sec. 4.4) that performs optimal one-to-one matching between LDLT and DDLT patients using all study variables listed in Sec. 4.2 (Fig. 1a). Unlike propensity score matching (PSM), which matches patients based on an output probability scalar, DPSM leverages and scores entire decision paths produced by a Random Forest model. This approach not only enhances the accuracy of the matching process but also increases the explainability of the method, providing a more transparent and interpretable framework for understanding factors influencing the matching.

**Fig. 1.**
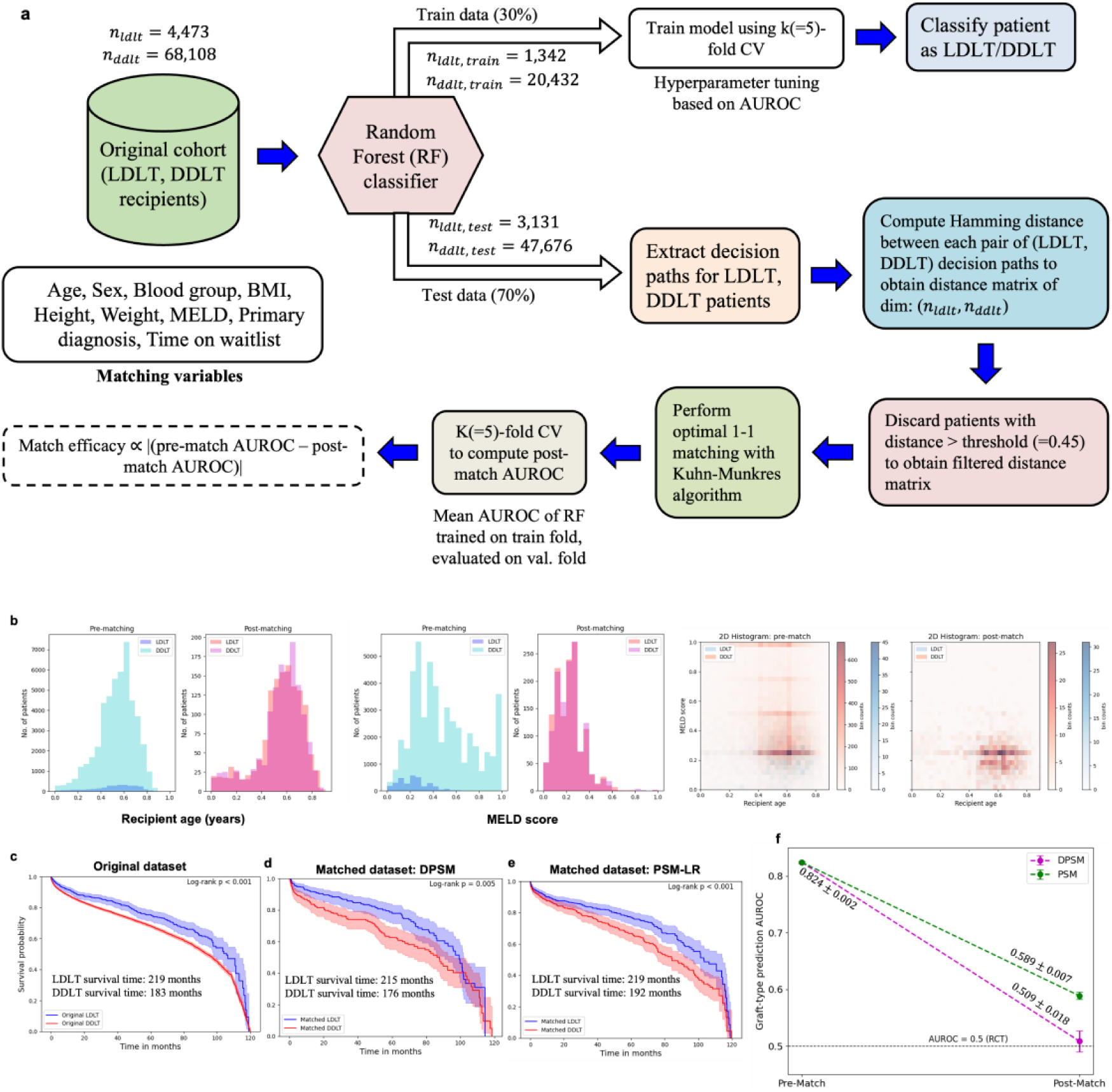
LDLT-DDLT Matching. (a) Workflow of our DPSM algorithm; (b) 1-D/2-D distributions (histogram) for recipient age and MELD score variables pre– and post-matching; (c) observed LDLT, DDLT survival in original dataset (pre-match); (d) observed LDLT, DDLT survival post-DPSM-matching; (e) observed LDLT, DDLT survival post-PSM-matching; (f) post-match AUROCs achieved by our DPSM and traditional PSM methods for the graft-type prediction task. The ideal case is AUROC=0.5, which corresponds to an actual RCT. Error bars indicate standard deviations across 10 random samplings.

First, we evaluated the effectiveness of our matching technique. Fig. 1b compares 1-D, 2-D pre– and post-match distributions for 2 key variables: age and MELD score across LDLT, DDLT patients. We observe a high degree of overlap for the matched populations, confirming the success of our method. Originally, DDLT patients had a relatively higher MELD score (22.4 ± 8.7) than those that received an LDLT (14.0 ± 4.5). High MELD (>33) patients, generally much sicker, are unable to be matched as they do not possess an LDLT counterpart. All other variables used in the study were also found to exhibit good matching (Fig. S2). Additionally, we sought to understand how matching impacts survival times of LDLT and DDLT patients. Kaplan-Meier analysis estimated LDLT and DDLT median survival times in the original dataset to be 219 and 184 months respectively (Fig. 1c). After matching using our DPSM technique, we observed a slight decrease in median survival times for the two groups (t_surv, ldlt_ = 215 months, t_surv, ddlt_ = 176 months) (Fig. 1d). For completeness, survival times after PSM matching were also computed (t_surv, ldlt_ = 219 months, t_surv, ddlt_ = 192 months) (Fig. 1e).

Next, we quantitatively evaluated the efficacy of our DPSM method by computing the AUROC of a Random Forest classifier trained on the pre– and post-matched datasets (Fig. 1f). The mean AUROC performance dropped significantly from 0.83 on the original dataset to 0.51 after matching, indicating that the model found it challenging to accurately classify patients as LDLT or DDLT, thus demonstrating the success of our matching process in reducing systematic differences between the two groups. Notably, post-match AUROC for DPSM (0.509 ± 0.018) was lower than that achieved using traditional PSM (0.589 ± 0.007), suggesting that our method performed better in achieving balanced and comparable groups. This enables establishment of a clearer causal relationship between graft type and survival outcomes.

A key advantage of our DPSM method is its ability to provide explainable matching, enhancing the transparency of the model’s decision-making process. To illustrate this, we show the 10 most frequently occurring patient variables in the decision paths across all trees of the Random Forest. This type of interpretability is essential because it transforms a complex, “black-box” setup into something that can be understood by researchers and clinicians. Fig. 2 shows these explanations for three randomly selected LDLT-matched DDLT patient pairs, where we observe that MELD score, waitlist time, weight and BMI are the top variables that influence the model deciding whether a patient received an LDLT or DDLT. Other patient characteristics such as age, sex and height were also found to be important predictors. Among etiologies, Alcoholic Cirrhosis, HCV Cirrhosis and PSC were deemed important by the Random Forest model to make graft-type predictions.

**Fig. 2.**
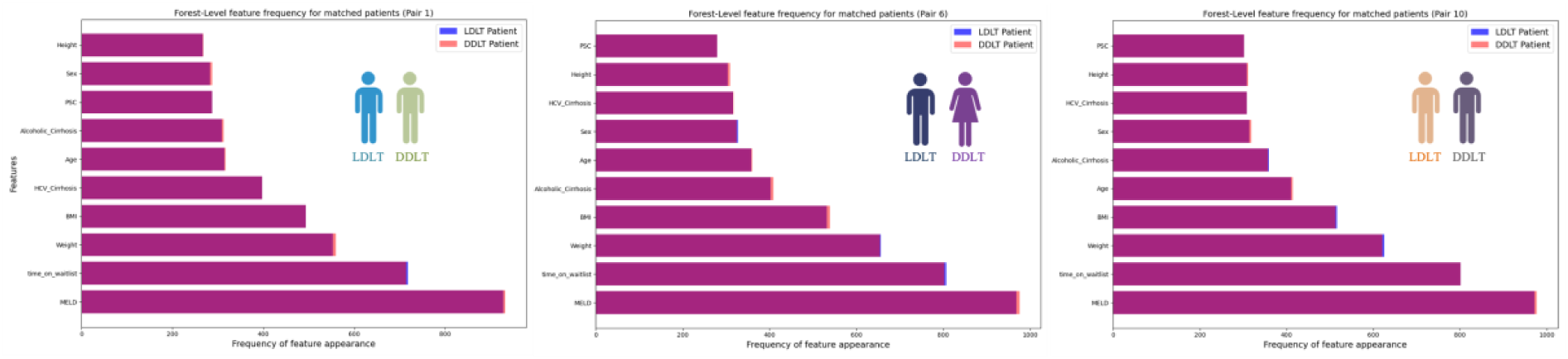
Frequently occurring patient characteristics across forest-level decision paths. For a given LDLT-matched DDLT patient pair, we show the 10 most frequently occurring variables across decision paths across all Trees of the Random Forest. A high degree of overlap indicates similarity between the two patients (differing by graft-type) in terms of how the forest makes decisions. This is illustrated for 3 randomly chosen patient pairs.

### 2.2. Evaluation of survival model

Building on the matched LDLT and DDLT cohorts generated by DPSM, we trained and evaluated survival models to predict patient survival outcome. Methodological details are provided in Sec. 4.5. For this task, we compared the performance of two popular time-to-event models, Random Survival Forest (RSF) and Cox Proportional Hazards (CPH). Two independent models were trained on the LDLT and matched DDLT populations.

In general, RSF (*C* − *index*_*ldlt*_ = 0.673, *C* − *index*_*ddlt*_ = 0.740) performed better than CPH (*C* −*index*_*ldlt*_ = 0.572, *C* − *index*_*ddlt*_ = 0.652) on the C-index and was therefore selected as the model of choice (Fig 3b). The improved performance may be attributed to the former’s ability to model non-linear data patterns without making explicit assumptions about underlying distributions. The average Brier score did not exceed 0.14. Additionally, we performed SHAP analysis to understand feature contributions to our outcome of interest, i.e., post-transplant mortality (Fig. 3c, d). Recipient age emerged as the strongest predictor of mortality with older patients at much greater risk. Other important factors were weight, BMI, MELD, height and blood type A. In terms of the primary indication for transplant, PSC, MASH, Alcoholic Cirrhosis and HCV Cirrhosis were all found to be important risk factors. In light of these findings, it becomes important to point out that variables such as age, MELD score, weight and BMI have been identified as key confounders. This is because these variables impact both, graft-type assignment (Fig. 2) as well as survival outcome.

**Fig. 3.**
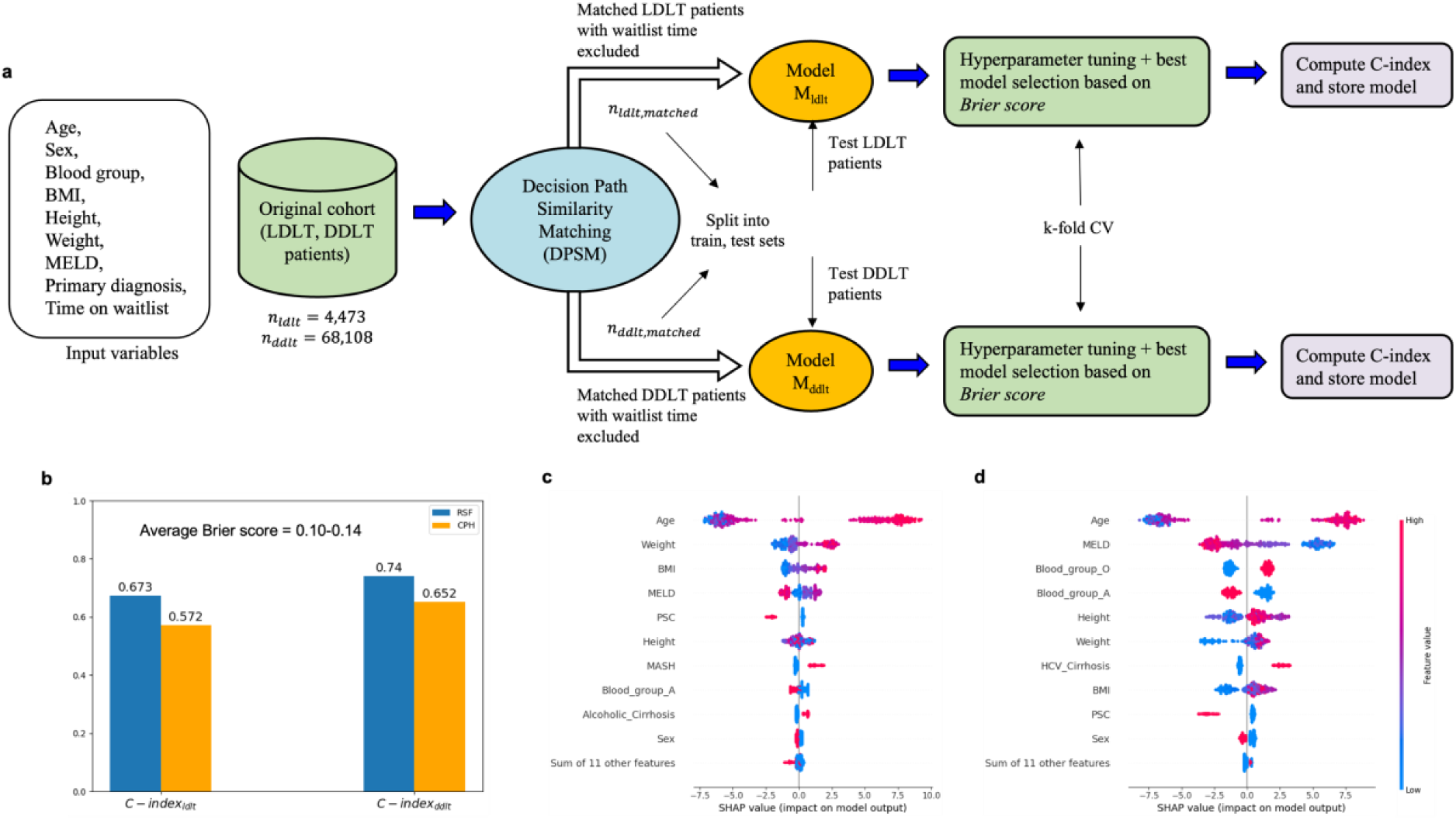
Performance evaluation and SHAP. (a) Model training and evaluation methodology: LDLT, DDLT cohorts are passed into the DPSM matching algorithm using all selected variables to account for confounding. Matched cohorts with waitlist time excluded are split into train and test sets. 2 graft-specific models are trained independently on the training samples, using a k-fold (k=5) cross-validation strategy and the best model across various hyperparameter settings is selected based on minimum Brier score. C-index is computed on the validation set and the best model is saved for further evaluation on the held-out test set; (b) RSF model C-index computed on the test set. Random Survival Forest (RSF) performance is compared with Cox proportional hazards (CPH) model, used as a baseline. Average Brier score varied from 0.10-0.14; top 10 features as predicted by SHAP on test set patients; (c) RSF-LDLT model applied to LDLT patients; (d) RSF-DDLT model applied to matched DDLT patients.

### 2.3. Estimation of LDLT benefit

A major strength of our ML-based technique is its ability to personalize survival prediction by incorporating individual patient variables into the modeling process. The emphasis in this section is on estimating the benefit in receiving an LDLT over a DDLT for an individual patient. To achieve this, we applied the trained RSF-LDLT and RSF-DDLT models to the LDLT and their matched DDLT counterparts, respectively. By contrasting the survival predictions generated by these models, we can estimate the personalized benefit of LDLT for each patient.

Fig. 4 shows individual examples for two etiologies of interest: for the patient diagnosed with Primary Sclerosing Cholangitis (PSC), our model predicts a 10-year survival gain of 16.8 % (Fig. 4b). To further understand the factors driving this predicted benefit, we conducted a patient-specific differential SHAP analysis (Fig. 4c), which identifies the key features contributing to survival gain. Variables highlighted in blue are associated with increased survival gain, while those in red indicate increased risk. The top variables that strongly influenced benefit were MELD, height, PSC and BMI. Original patient variables are also shown (Fig. 4d).

**Fig. 4.**
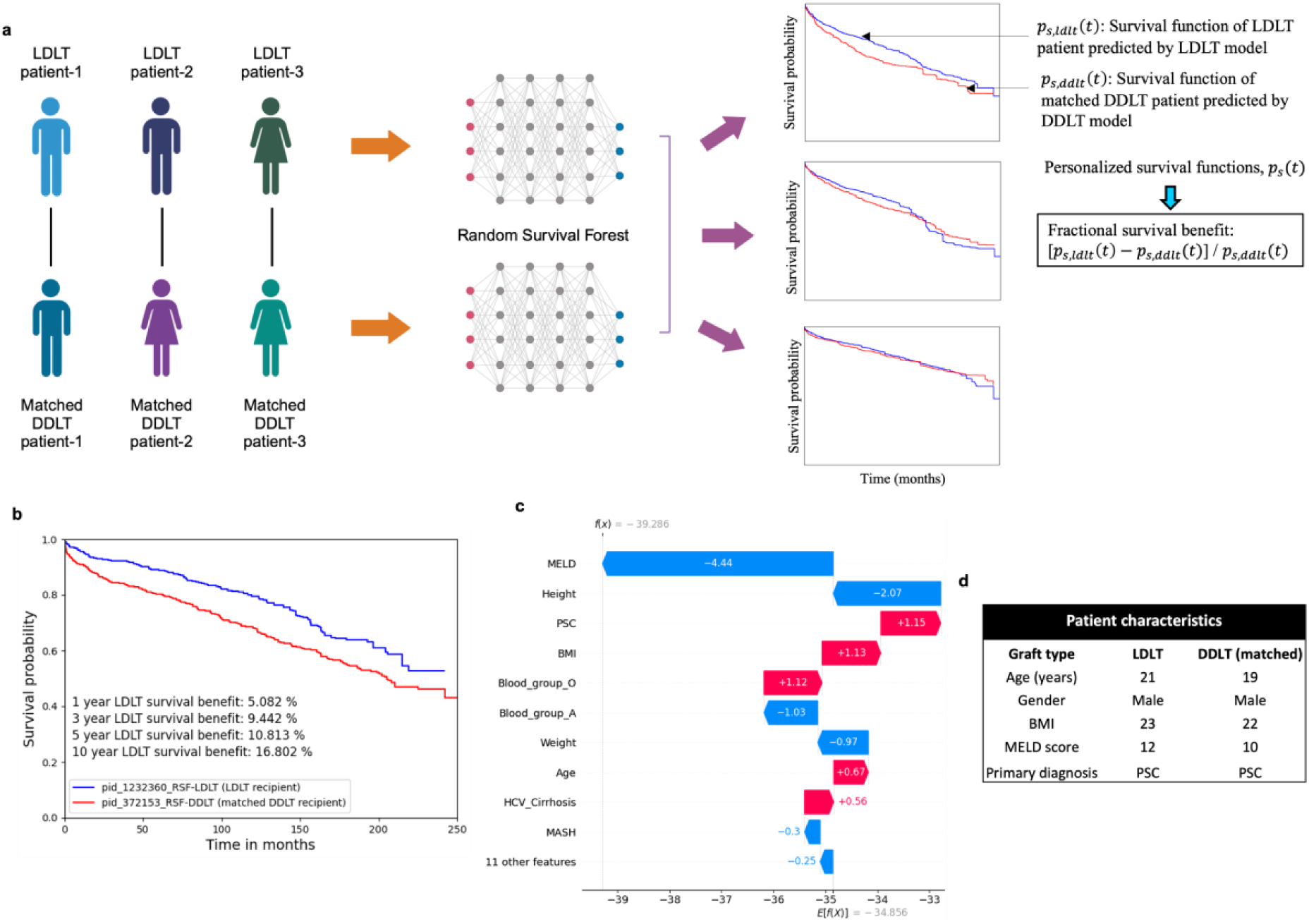
LDLT versus DDLT survival for individual patient. (a) Estimation of LDLT survival benefit: LDLT, DDLT survival models are applied independently to obtain the predicted survival functions p_s, ldlt_(*t*) and p_s, ddlt_(*t*)for LDLT and matched DDLT recipients (test), respectively. At a desired evaluation time-point, fractional LDLT survival benefit is defined as the probability difference between LDLT and DDLT survival normalized by DDLT survival; (b) LDLT benefit (%) at 1-, 3-, 5– and 10-years post-transplant for a patient diagnosed with PSC. Blue and red curves show survival functions of the LDLT and matched DDLT patient predicted by the respective models. Individual SHAP explanations (c) as well as corresponding patient characteristics (d) are also shown.

Next, we compute the population-level predicted LDLT benefit by considering the matched patient groups. This is done on the held-out test set of patients, untouched during the model training process (Fig. 5a). Based on variables at the time of listing, our ML model predicts a mean long-term (10-year) benefit of 10.3%. For validation, we also performed standard Kaplan-Meier analysis on the same set of patients to evaluate observed LDLT versus DDLT survival differences (Fig. 5b).

**Fig. 5.**
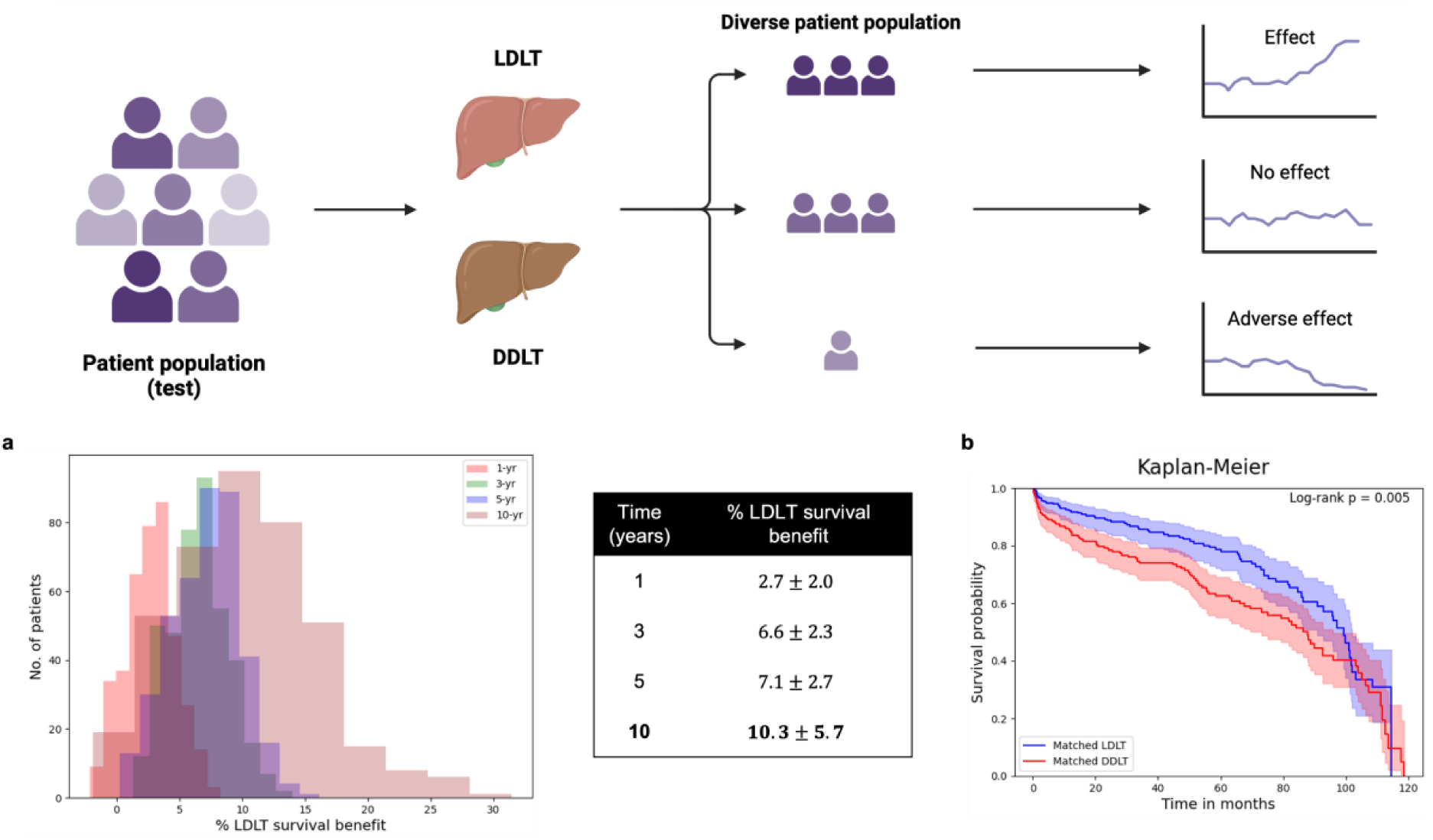
LDLT survival benefit. (a) 1, 3, 5 and 10-year fractional survival benefit of receiving LDLT over DDLT (*n*_*test*_ = 401) for LDLT recipients. If *t* does not exist for either of the LDLT, DDLT models, we perform interpolation, so that % benefit can be computed appropriately; (b) Kaplan-Meier estimator applied on the test set confirms LDLT survival benefit at the population-level. Survival times are cut-off at 120 months (10-years).

### 2.4. Etiology-specific benefit

Finally, we evaluated the predicted survival benefit of LDLT over DDLT across six different etiologies (primary diagnoses) within the matched cohort, namely Autoimmune Hepatitis (AH), PBC, PSC, HCV Cirrhosis (HCV), MASH, and Alcoholic Cirrhosis (AC). By analyzing the survival outcomes for specific diseases, we aimed to identify which etiologies were associated with the highest LDLT benefit.

Using our ML-guided approach, we computed the differential survival gain of LDLT for each patient and then aggregated these results based on their primary diagnosis (Fig. 6a). The analysis revealed that certain etiologies exhibited a significantly higher survival benefit when transplanted with LDLT compared to DDLT. Patients diagnosed with PSC (12.4 ± 5.3 %) and HCV (12.1 ± 5.7 %) showed substantial long-term survival advantages with LDLT, over a 10-year period. For comparison with ground truth, we also evaluated observed survival differences between the two groups (Fig. 6b). These findings suggest that LDLT may be particularly advantageous for patients with these conditions, potentially influencing clinical decision-making.

**Fig. 6.**
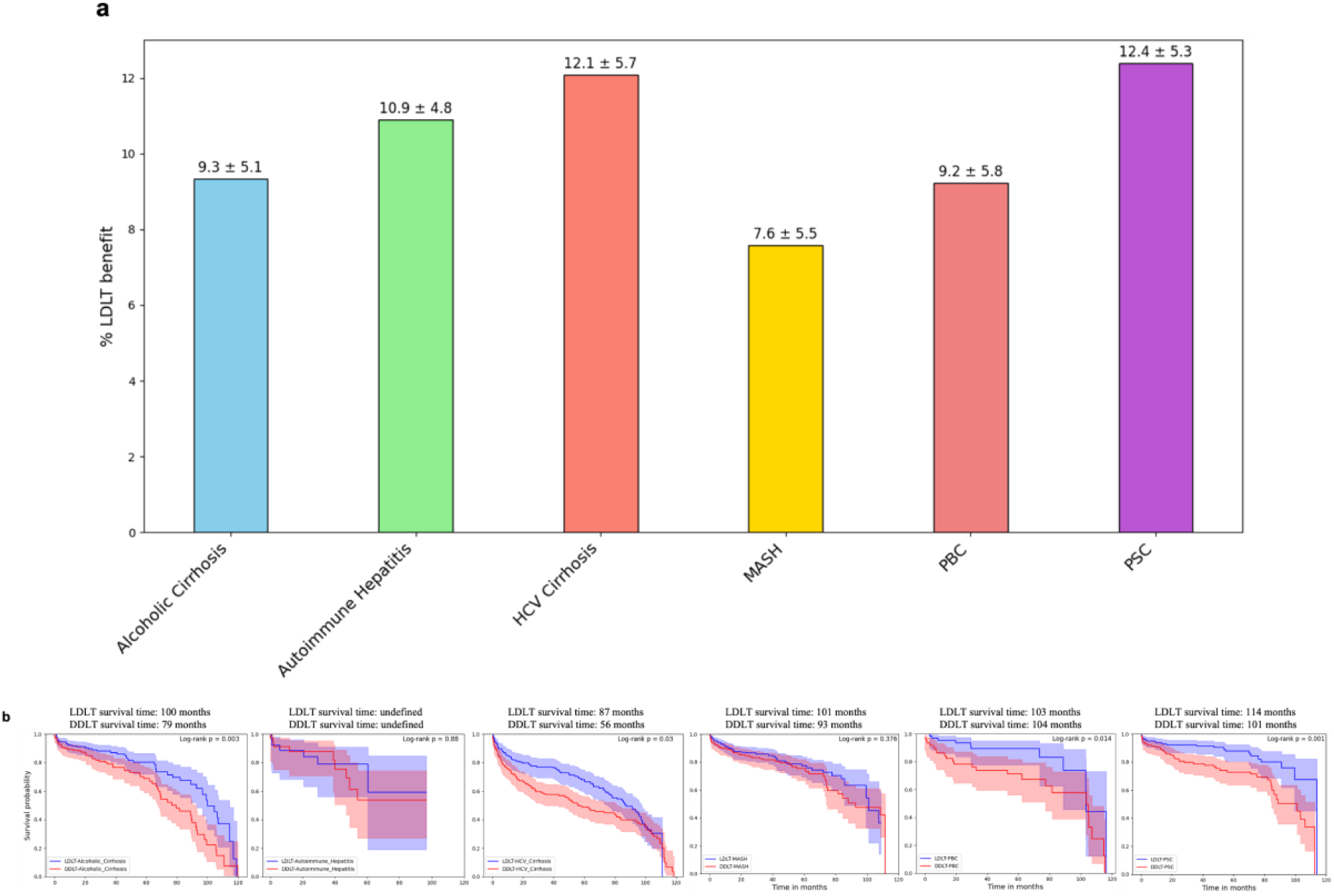
Etiology-specific benefit. (b) Bar chart shows 10-year (long-term) post-transplant LDLT benefit (%) for each of the 6 distinct etiologies: Alcoholic Cirrhosis, Autoimmune Hepatitis, HCV Cirrhosis, MASH, PBC and PSC. Results are computed on the held-out test set, these patients are untouched during RSF model training; (b) observed survival (Kaplan-Meier) for all 6 etiologies to evaluate matched LDLT (blue) vs DDLT (red) survival differences. Survival data is cut-off at 10 years.

## 3. Discussion

The challenge of determining which patients would benefit most from receiving a living donor liver transplant (LDLT) versus a deceased donor liver transplant (DDLT) is compounded by the impracticality of conducting randomized controlled trials (RCTs). To address this, we create a novel approach which we call Decision Path Similarity Matching (DPSM) algorithm, that combines advanced matching and machine learning (ML) techniques to emulate the balanced conditions of an RCT using observational data to personalize survival predictions. By effectively minimizing confounding factors, DPSM enables causal-type estimation of the effect of graft-type on post-transplant survival.

### 3.1. DPSM: a novel, multivariate method for one-to-one matching

Our innovative algorithm represents a significant advancement in the field of observational study design, particularly in contexts where conducting randomized controlled trials (RCTs) is impractical or impossible. Unlike traditional Propensity Score Matching (PSM), which compresses complex patient data into a single propensity score, DPSM leverages the full decision paths generated by a Random Forest classifier to perform one-to-one matching between LDLT and DDLT patients. This approach allows DPSM to retain the multidimensional complexity of patient data, resulting in more nuanced and accurate matching that closely mimics the balance achieved in RCTs.

The key strength of DPSM is its ability to minimize confounder bias, thereby producing more balanced cohorts than traditional PSM. Our study demonstrated that DPSM significantly decreased the AUROC for graft-type prediction after matching, a clear indication that DPSM more effectively mitigates covariate differences between the LDLT and DDLT groups. This enhanced performance is crucial in creating a robust foundation for subsequent survival analysis, ensuring that any observed differences in outcomes are attributable to graft-type differences.

Additionally, by utilizing decision paths instead of a single scalar score, DPSM allows clinicians and researchers to understand the prominent variables involved in the decision-making process leading up to the matching. These insights are particularly valuable in clinical settings, where understanding the rationale behind matching decisions can foster greater confidence in the study’s findings and support more informed clinical decision-making. The ability to visualize and interpret the decision-making process also aligns DPSM more closely with the principles of RCTs, where the reasoning behind patient assignment is clear and systematic.

In summary, DPSM is a powerful technique, enabling researchers to simulate the conditions of an RCT more effectively in observational studies. Its superior matching performance and transparency make it a valuable tool not only for advancing research in liver transplantation but also for broader applications where the target questions are causal in nature. As the field of clinical research increasingly turns to observational data in the absence of feasible RCTs, methods like DPSM will play a critical role in ensuring that the insights drawn from these studies are both accurate and actionable.

### 3.2. ML framework for personalized survival predictions

Applying our RSF model on a test set of patients, we report a mean predicted LDLT survival gain of 2.7%, 6.6%, 7.1% and 10.3% at 1-, 3-, 5– and 10-years post-transplant respectively. It is worth placing this in the context of prior evidence pertaining to LDLT versus DDLT survival. Barbetta *et. al.*^10^ also analyzed SRTR data and found a mortality risk reduction of 17%, 15% and 13% at 1-, 3– and 5-year post-transplant for LDLT recipients. Higher estimated benefit in the latter is suspected to be a consequence of the lack of matching in their analysis. In fact, another study^11^ around the same time that analyzed Canadian transplant recipients found that graft-type differences got washed away upon adjustment for donor as well as recipient characteristics.

In our study, where we predict individual LDLT benefit using patient-specific variables at the time of listing, we find significant heterogeneity across all patients that we evaluated our method on, underscoring the importance of personalization. The ability to predict individualized survival outcomes allows clinicians to move beyond a ‘one-size-fits-all’ approach, enabling more tailored decisions that align with the specific characteristics and needs of each patient. This not only optimizes transplant outcomes but also enhances patient counseling, as clinicians can provide more accurate, data-driven information to patients and their families when discussing treatment options.

Finally, our etiology-specific analysis underscores the importance of individualized treatment planning, as the survival benefit of LDLT can vary significantly depending on the underlying liver disease. These insights could guide clinicians in making more informed decisions about transplant strategies, particularly for patients with specific diagnoses where LDLT offers the most substantial benefit. Among all etiologies tested, our ML tool predicted the highest benefit for patients diagnosed with PSC (12.4 ± 5.3 %). These patients often experience slow progression of the disease, but when complications such as cholangitis arise, timely transplantation becomes critical. LDLT, with its shorter wait times, offers a significant survival advantage in such urgent cases. Our result is backed by a recent study by Sierra *et. al.*^17^, which also reported a long-term (10-year) survival advantage for PSC patients who received an LDLT (81.9 %) over a DDLT (72.7 %).

Integrating these personalized predictions into clinical workflows could significantly improve decision-making processes, ensuring that patients are referred for LDLT especially when the survival benefit is significant. As the field of liver transplantation progresses towards precision-based approaches, tools like these will become essential in guiding downstream clinical decisions and potentially improving overall patient care.

### 3.3. Study strengths and limitations

This study has several notable strengths that enhance its contribution to the field of liver transplantation. First, the development and application of the Decision Path Similarity Matching (DPSM) algorithm represents a significant advancement in observational study design, allowing us to create well-matched cohorts that closely mimic the conditions of an RCT. This is evident in the substantial reduction in systematic differences between the LDLT and DDLT cohorts, as evidenced by the drop in AUROC from 0.83 (pre-matching) to 0.51 (post-matching), which shows that our algorithm effectively removed confounding associations among variables. By reducing bias and aligning survival outcomes across groups, DPSM enables a more accurate comparison of LDLT and DDLT outcomes, which is crucial in the absence of feasible RCTs. Additionally, the integration of Random Survival Forest (RSF) models to provide personalized survival predictions adds a valuable dimension to clinical decision-making, offering tailored insights that can optimize patient care.

Our study also has important limitations. The retrospective nature of the analysis, relying on data from the SRTR, may introduce biases inherent in observational studies. It is important to note that DPSM is able to mitigate these confounding biases, as long as they are observable, i.e., captured within our dataset. Another limitation is the reliance on clinico-demographic variables available at the time of listing, which, although comprehensive, may not capture all factors influencing transplant outcomes. Incorporating additional variables, such as genetic markers or more detailed comorbidity data, could further refine the predictive models and enhance the precision of survival estimates.

We also acknowledge the absence of external validation as a limitation of this study. This was primarily due to the lack of access to sufficiently large datasets outside the SRTR. DPSM performs optimally with larger sample sizes, especially the DDLT pool, as these allow for more reliable transformation of observational data into RCT-like conditions. Smaller datasets may not provide the robustness needed for effective matching, underscoring the importance of future research focused on validating these findings in different cohorts and clinical environments. Finally, while the RSF model demonstrated good performance in this study, further validation in prospective studies and across different cohorts is necessary to confirm its broader applicability.

In conclusion, this study introduces a novel Decision Path Similarity Matching (DPSM) methodology, which represents a significant advancement in creating RCT-like comparability from observational data and debiasing transplant outcomes. DPSM offers a more transparent and explainable approach to matching patients compared to traditional methods, allowing for personalized predictions of survival benefit in living donor liver transplantation. While further research and external validation are required to enhance its robustness and generalizability, the innovations presented here mark an important step toward more individualized, data-driven decision-making in liver transplantation.

## 4. Methods

### 4.1. Study Design

We conducted a retrospective, cross-sectional study from the SRTR database. The SRTR includes data of all organ donors, as well as waitlisted and transplanted recipients in the United States submitted by the Organ Procurement and Transplantation Network (OPTN). This study was approved by the Research Ethics Board at the University Health Network.

Adult (≥18-years-old) LT patients, listed between 28^th^ February 2002 and 23^rd^ May 2023, were included in the analysis. Study exclusion criteria is clearly defined in Fig. S1. Patients with reported MELD scores greater than 40 were excluded. These patients (<5% of the entire population) are generally very sick and highly prone to pre-operative mortality, including them would make it challenging to clearly delineate the effect of transplant type on post-transplant survival. Recipients with previous or multi-organ transplants were excluded as were those diagnosed with HIV, acute liver failure (ALF) and HCC. Patients who received exception points were also removed.

### 4.1. Variables and Outcomes

Clinico-demographic patient variables: age, sex, blood group, BMI, height, weight, MELD score, primary diagnosis (indication for transplant) and time on the waitlist were collected at the time of listing. For variable comparison between the LDLT, DDLT patient cohorts, an alpha level of <0.05 was selected as the significance threshold. Post-transplant, mortality (all-cause) was the outcome or event of interest and event times were defined from the time of transplant to either the time of death or censored at the date of last follow-up.

### 4.2. Data Preparation and Preprocessing

Covariates with greater than 20% missingness were excluded from the analysis. For continuous variables with missing values, mean imputation was performed. Categorical variables were one-hot encoded (OHE). Subsequently, we ended up with 21, unit normalized input features to the ML model.

### 4.3. Matching

We designed and implemented a new method, Decision Path Similarity Matching (DPSM) to account for confounding bias, ensuring that predicted survival differences would be predominantly attributed to graft-type. The key feature of DPSM being that it uses the “similarity” or closeness between decision paths, which provides a richer feature encoding as opposed to matching based on output probabilities alone in the case of PSM. The constituent steps of our algorithm are shown in Fig. 1a along with the pseudocode below (Fig. 7) – (1) first, the original LDLT-DDLT dataset is randomly split into train (70%) and test (30%) sets. A Random Forest (RF) classifier is trained on the training set to predict transplant type using all input variables previously listed in Sec. 4.2. RF was selected due to its ability to capture non-linear relationships among variables and handle imbalanced data, crucial in the LDLT versus DDLT context. (2) The best model is selected across a hyperparameter search (*n*_*estimato*r*s*_ = 50, 100, 200, 300, 500, 1000, *min*_*samples*,*leaf*_ = 500) using k(=5)-fold cross-validation (cv) on the train set and performance is evaluated on the test set using the area under the receiver operator characteristic (AUROC) curve. (3) From the trained model, we extract decision paths for individual patients in the held-out test set, averaged across all the Trees in the Forest. This is an n-dimensional binary vector [1, 0, 1, 0,.], where “n” is the total number of decision nodes (features) per tree. A node is “1” if it applies to a particular patient and “0” otherwise. (4) Hamming distance (*d*_*H*_) is then computed for every pair of (LDLT, DDLT) decision paths. To remove “poor” matches or outliers, patients whose pairwise *d*_*H*_ exceeds a selected threshold (*t*ℎ) are removed. The optimal threshold was determined as that which minimized |*AU*R*OC*_*post*−*matc*ℎ_ − 0.5|, at an acceptable patient dropout rate (Table S1). We selected *t*ℎ = 0.45. (5) The filtered distance matrix is used to perform one-to-one matching using the “Munkres Assignment” procedure^18^. This is an “optimal” algorithm that matches by minimizing the total Hamming distance across all LDLT-DDLT patient pairs. After this step, we repeat step (2) to calculate the AUROC for the matched dataset using a k(=5)-fold cv strategy. The same set of hyperparameters are used. Finally, match effectiveness is quantified as the difference between pre– and post-match AUROCs.

**Fig. 7.**
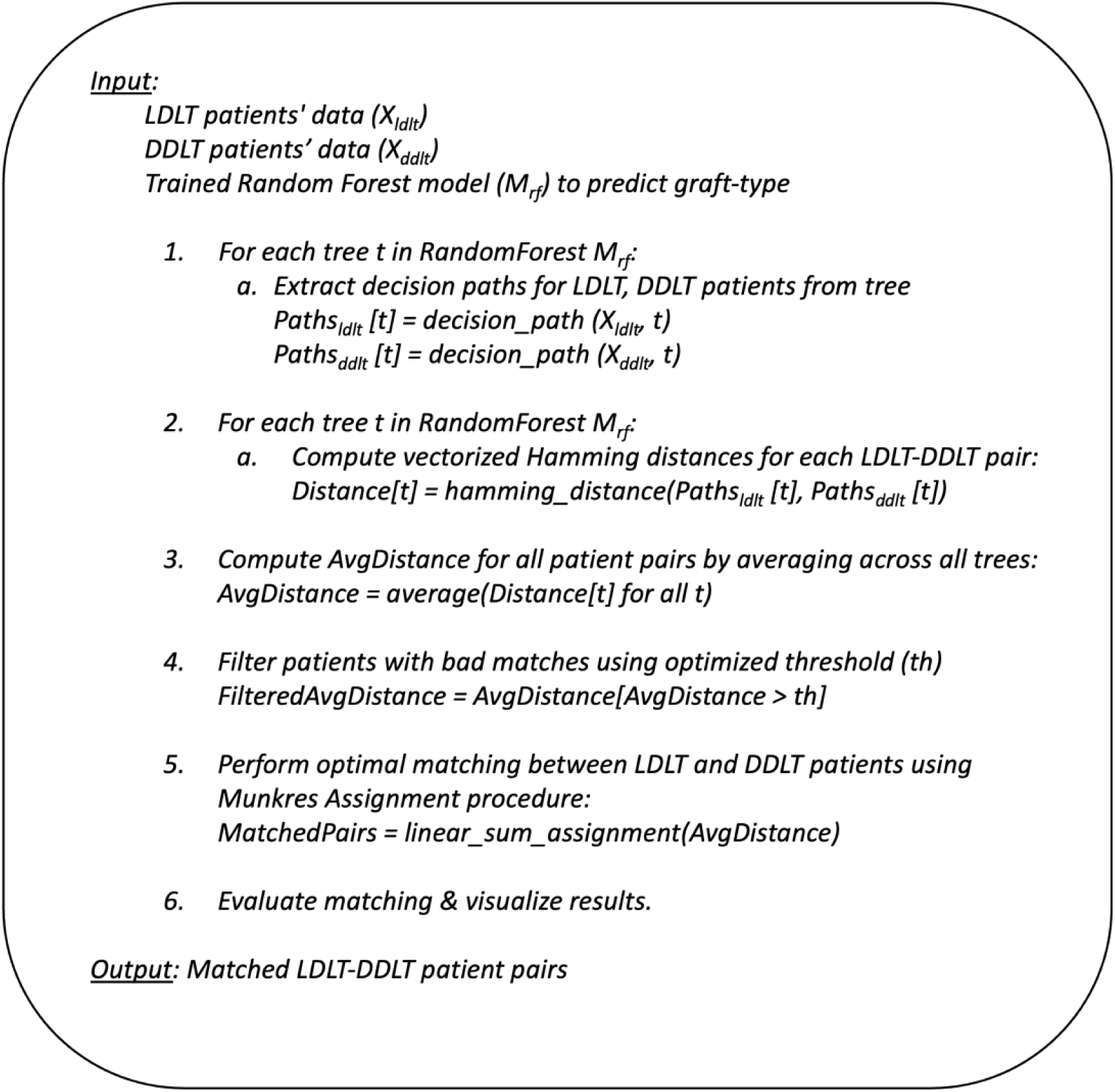
DPSM: pseudocode.

### 4.4. Model Training and Validation

The matched sub-populations (*n*_*ldlt*_ = *n*_*ddlt*_ = 1,337) were first split into train (70%) and test (30%) sets (*n*_*t*r*ain*_ = 936, *n*_*test*_ = 401). For the time-to-event survival prediction, two Random Survival Forest (RSF) models were trained independently on the matched LDLT-DDLT cohorts, respectively. Hyperparameter tuning was performed on the train dataset using k (=5)-fold cross validation and the model that produced minimum Brier score was selected as the best one. For validation, both C-index and Brier score, averaged across 5 evaluation time points: 0.5, 1, 3, 5 and 10 years were computed on the held-out test set.

During hyperparameter tuning, it is common to use C-index as the evaluation metric for time-to-event models. While this is useful in understanding relative risk ranking, it is not informative about the accuracy and calibration of the predicted survival predictions. We instead utilize the Brier score, defined as follows: 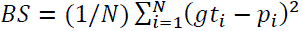, where N is the sample size, *gt*_*i*_ and *p*_*i*_ are the actual and predicted event probabilities for observation *i*. This metric quantifies the error rate between prediction and ground truth, serving as an ideal choice for calibrating survival models. Subsequently, the optimal hyperparameters selected are those that minimize the time-averaged Brier score.

### 4.5. Estimation of LDLT Survival Benefit

For a given LDLT and matched DDLT patient, the corresponding trained models (RSF-LDLT, RSF-DDLT) are applied to obtain the respective predicted survival functions *S*_*ldlt*_ (*t*) and *S*_*ddlt*_(*t*). RSF models produce unique event times according to the data they were trained on. To ensure that both LDLT and DDLT models predict definitive survival for a given evaluation time point *t*_*i*_, we interpolate predicted survival probabilities across all time. Finally, differential LDLT benefit is evaluated as (*S*_*ldlt*_ (*t*) − *S*_*matc*ℎ*ed ddlt*_(*t*))/*S*_*matc*ℎ*ed ddlt*_ (*t*).

## 5. Code availability

The source code for this work is available on GitHub (https://github.com/Anivader/LDLT_survival_benefit_ML_tool). All analysis was performed using Python.

## Data Availability

All data produced in the present study are publicly available through the Scientific Registry of Transplant Recipients (SRTR) and available upon reasonable request to the authors. The source code for this work is available on GitHub.

https://github.com/Anivader/LDLT_survival_benefit_ML_tool

## 7. Acknowledgements

Mamatha Bhat acknowledges support from the Toronto General and Western Hospital Foundation, Canadian Institutes for Health Research and Canadian Donation and Transplant Research Program. Chris McIntosh holds the Chair in Medical Imaging at the Joint Department of Medical Imaging at the University Health Network, and the Department of Medical Imaging at the University of Toronto. Michael Brudno holds a CIFAR AI Chair.

## 8. Author contributions

A.G., C.M., M. Bhat. and Y.S. conceptualized the study. C.M. supervised the experimental design, A.G. developed the computational analysis pipelines and generated all the data. Y.S. helped with the data pre-processing script. B.J.H. and X.Z. helped with clinical interpretability. A.G. wrote the manuscript and C.M., B.J.H., M. Bhat provided feedback and M. Brudno reviewed the manuscript. C.M. and M. Bhat. supervised the study.

## 9. Ethics declarations

The authors declare no competing interests.

