## Supplemental Materials for "A confounder debiasing method for RCT-like comparability enables Machine Learning-based personalization of survival benefit in living donor liver transplantation"

### Supplementary Information

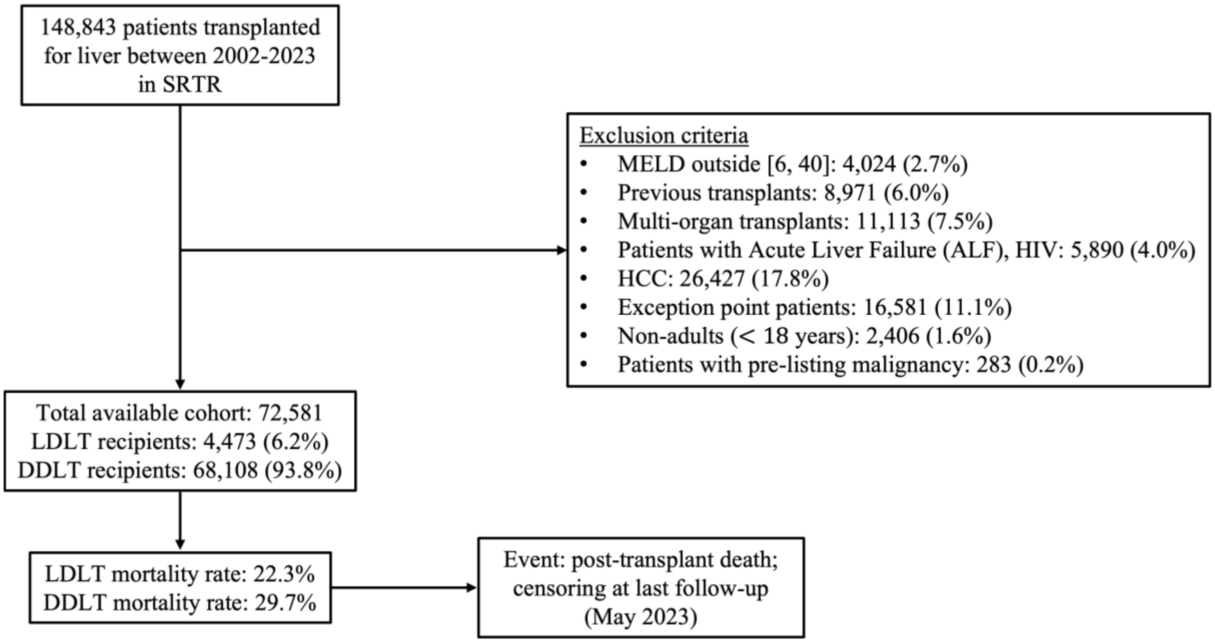


**Fig. S1. Patient inclusion-exclusion criteria.**

<table>
<tr><th>$N_{LDLT, original}$</th><th>Threshold</th><th>$N_{LDLT, matched}$</th><th>$AUROC_{pre\text{-}match}$</th><th>$AUROC_{post\text{-}match}$</th></tr>
<tr><td rowspan="5">3,131</td><td>1</td><td>2,818 (90%)</td><td rowspan="5">0.825</td><td>$0.567 \pm 0.014$</td></tr>
<tr><td>0.75</td><td>2,372 (76%)</td><td>$0.559 \pm 0.020$</td></tr>
<tr><td>0.5</td><td>1,575 (50%)</td><td>$0.537 \pm 0.021$</td></tr>
<tr><td>0.45</td><td>1,337 (43%)</td><td>$\mathbf{0.518 \pm 0.013}$</td></tr>
<tr><td>0.4</td><td>1,089 (35%)</td><td>$0.497 \pm 0.014$</td></tr>
</table>

**Table S1. Effect of filtering threshold on matching performance.**
Our DPSM method removes patients with bad matches by applying a filtering threshold to the computed distance matrix. LDLT and DDLT patients with pairwise average Hamming distance greater than the threshold are removed from the dataset prior to matching. The optimal threshold was determined as that yielding a post-match AUROC ~ 0.5, while keeping the patient dropout rates reasonable. We selected $thresh = 0.45$.

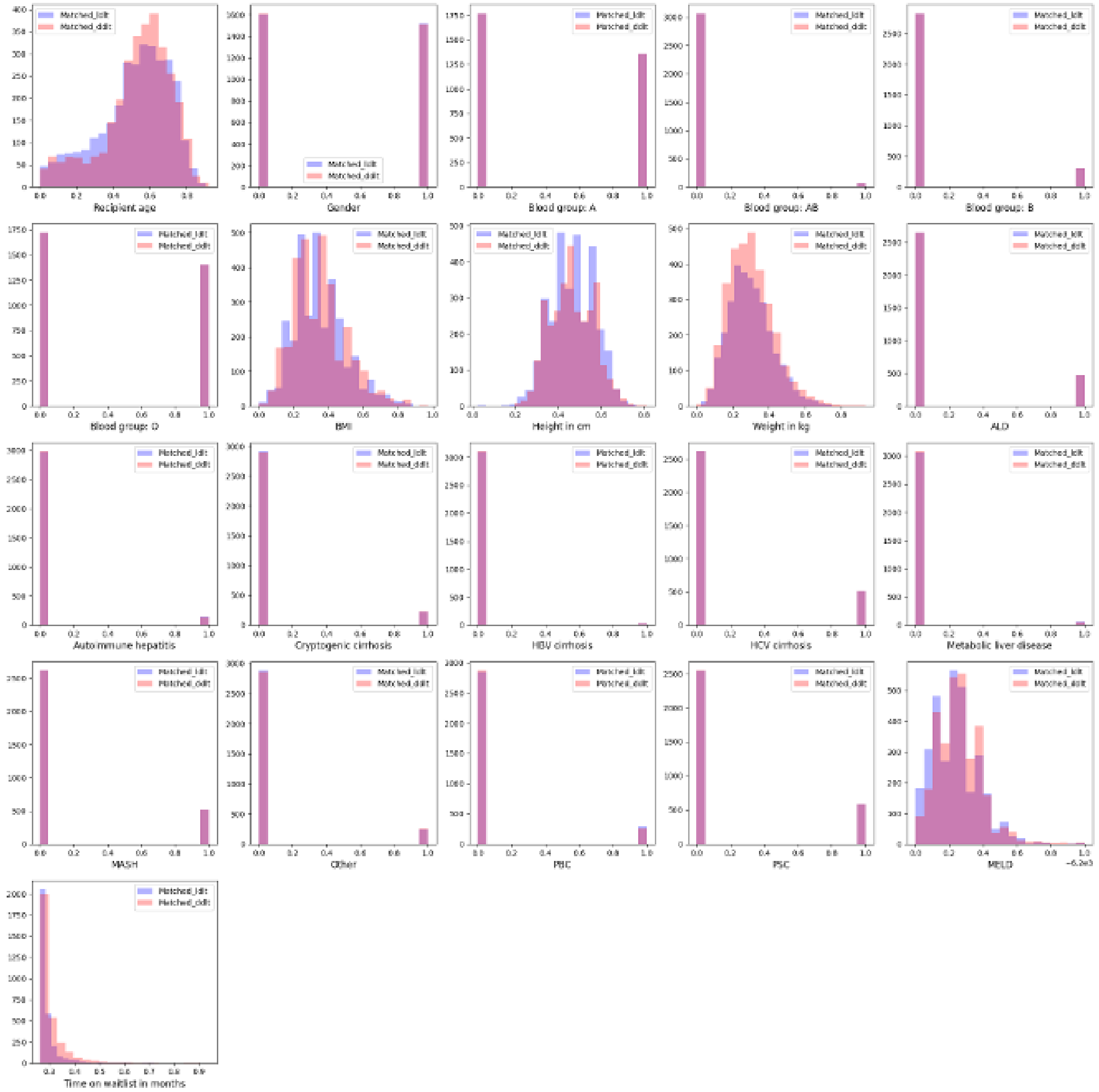


**Fig. S2. Covariate distributions post-matching.**
Abscissa shows normalized covariate values.

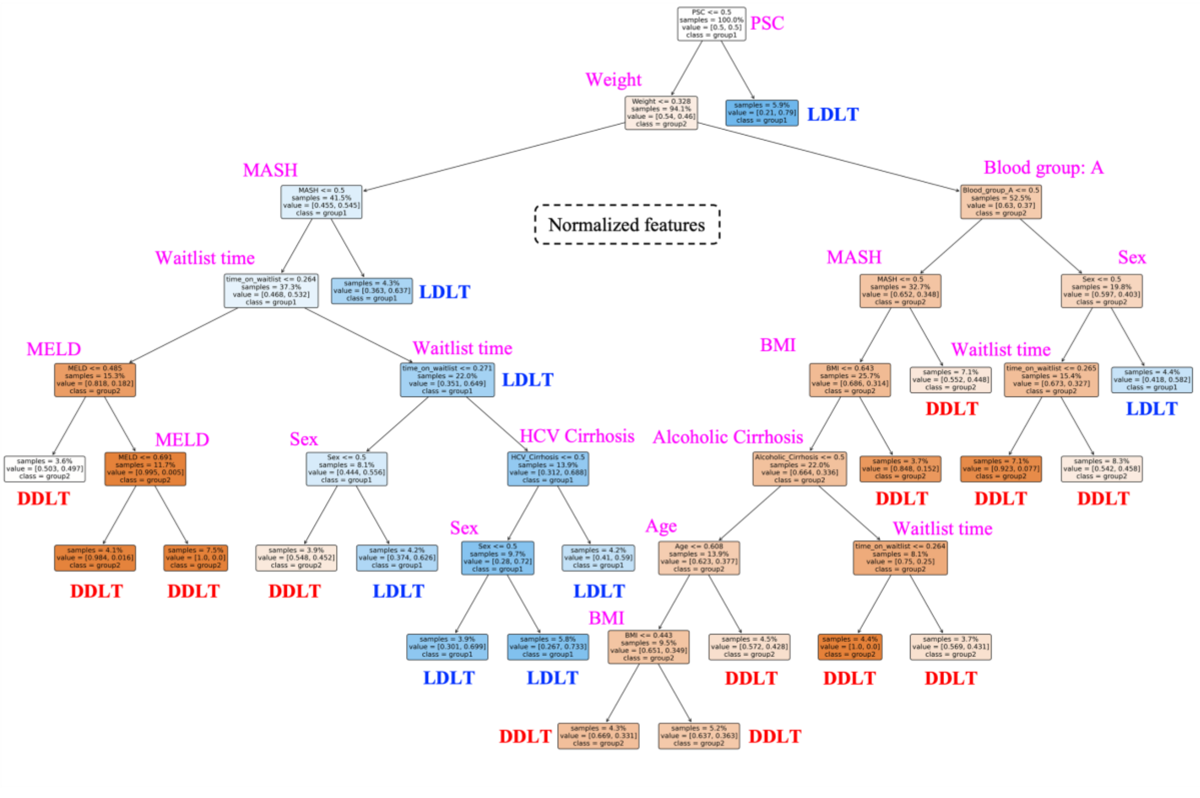


**Fig. S3. Decision path of a single tree**

Decision path of a single Tree in the "Forest". This informs as to which features were prioritized and used in making graft-type predictions. Features are normalized.

7
